# Twelve-Month Follow-Up of Patients With Generalized Myasthenia Gravis Receiving BCMA-Directed mRNA Cell Therapy

**DOI:** 10.1101/2024.01.03.24300770

**Authors:** Nizar Chahin, Gregory Sahagian, Marc H. Feinberg, C. Andrew Stewart, Christopher M. Jewell, Metin Kurtoglu, Miloš D. Miljković, Tuan Vu, Tahseen Mozaffar, James F. Howard

## Abstract

We report the 12-month follow-up results of a phase 2 clinical of Descartes-08 (NCT04146051), BCMA-directed RNA chimeric antigen receptor T-cell (rCAR-T) therapy for myasthenia gravis (MG) given as an outpatient treatment without lymphodepletion. In the Phase 2a part of the study, all 7 participants who received six weekly infusions of Descartes-08 exhibited clinically meaningful improvement in common MG severity scales (MG Composite, MG Activities of Daily Living, Quantitative MG scores, and Quality of Life 15-revised) at Month 3. At Month 9 follow-up, all participants continued to experience marked clinical improvements. Five out of seven participants maintained clinical improvement at Month 12. Of the two participants who experienced loss of clinical effect at Month 12 and were eligible for retreatment, one was retreated and had rapid improvement in clinical scores with minimal symptom expression which was ongoing at Month 6 of follow-up. All three participants with detectable anti-acetylcholine receptor (AChR) antibody levels at baseline experienced autoantibody reductions by Month 6, which deepened further by Month 9, and were maintained at Month 12. These data support continued development of Descartes-08 in myasthenia gravis and other autoantibody-associated autoimmune disorders.

---

We recently reported on the safety and preliminary clinical activity of Descartes-08, an mRNA-engineered chimeric antigen receptor (rCAR) T-cell therapy, in patients with generalized myasthenia gravis (gMG) (NCT04146051).^1^ Descartes-08 is administered in the outpatient setting, does not require lymphodepleting chemotherapy, and targets B-cell Maturation Antigen (BCMA) expressed on long-lived plasma cells (LLPCs) and plasmacytoid dendritic cells (pDCs).^2^ In the Phase 2a part of the study, all 7 participants (4 acetylcholine receptor [AChR]+, 2 muscle-specific kinase [MuSK]+, 1 seronegative, by history) who received six weekly infusions of Descartes-08 exhibited clinically meaningful improvement in common MG severity scales at Month 3. Three of the 7 participants achieved minimal symptom expression (MSE)^3^. Two were intravenous immunoglobulin (IVIG) infusion-dependent and one was plasmapheresis-dependent for years prior to treatment, and maintained responses without further IVIG or plasmapheresis. Here, we report the results of the final per-protocol one year follow-up.

Clinical trial design, objectives, outcome measures, laboratory and data analysis methods, and demographic characteristics were previously described.^1^ Descartes-08 maintained a favorable safety profile, with no new product-related adverse events reported during the 12-month follow-up period. At Month 9 follow-up, approximately 7 months after the last infusion and without new or increased MG-directed drugs, all participants continued to experience substantial clinical responses as measured by the MG Activities of Daily Living Score^4^ (mean change -6.3 [95% CI -3·5 to -9.1], **Figure 1A**), MG Composite Score^5^ (−16·6 [-13·4 to -19·5, **Figure 1B**), Quantitative MG scores^6^ (−8·4 [-5·2 to -11·6], **Figure 1C**), and Quality of Life 15-revised score^7^ (−12 [-8 to -16], **Figure s1**). At Month 12, 5 of the 7 participants maintained clinically meaningful improvement, including one with MSE. Two participants had worsening disease at Month 12 and became eligible for retreatment. One opted for retreatment, which led to a rapid decrease in MG-specific clinical scores and MSE. The MSE is ongoing at Month 6 of retreatment follow-up (**Figure 1D**).

**Figure 1.**
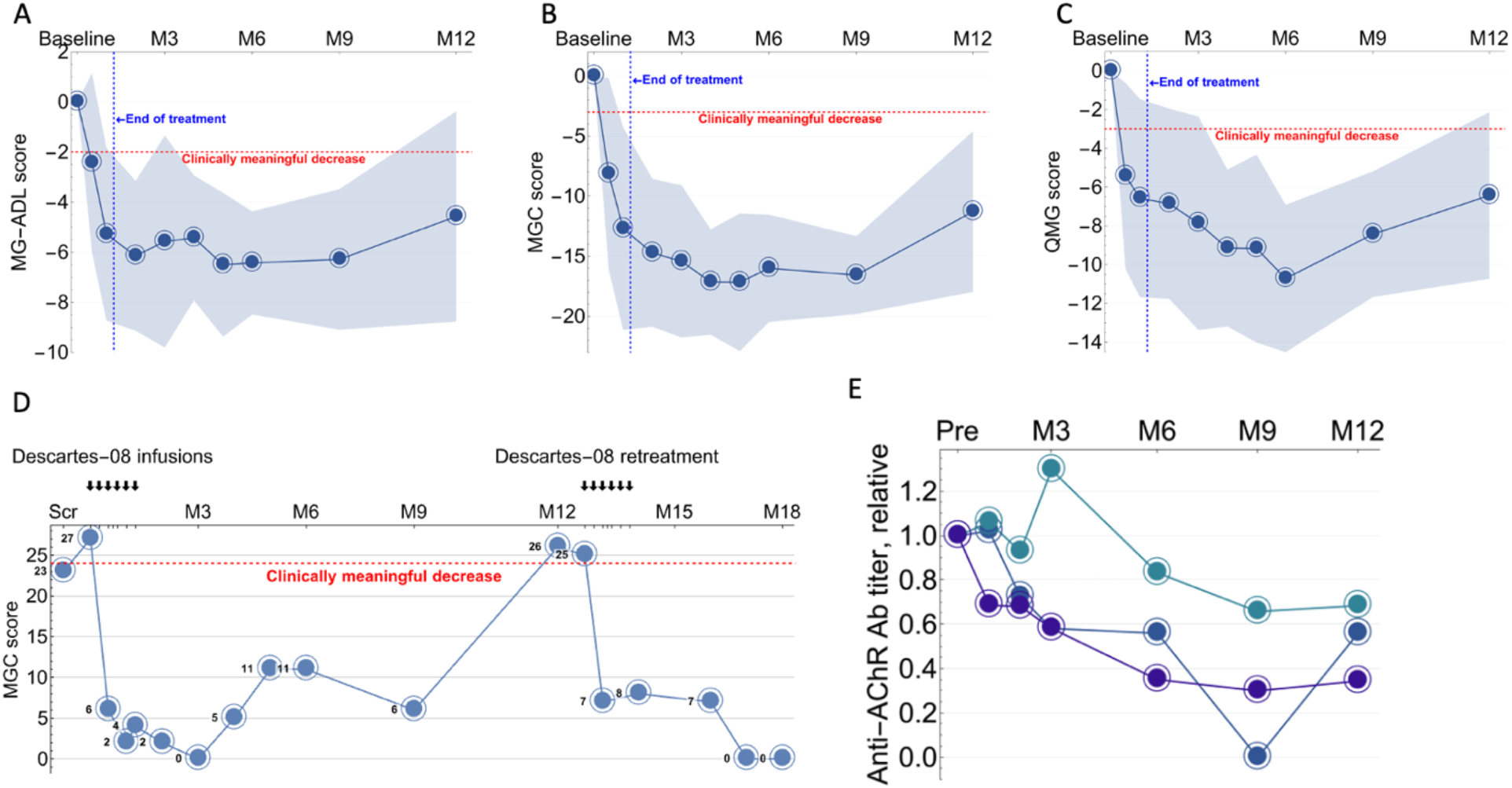
Changes in disease severity scores and serum autoantibody levels. **A–C:** Mean change from Baseline (line) and 95% CI (bands) in Myasthenia Gravis Activities of Daily Living Score (MG-ADL, **A**), Quantitative Myasthenia Gravis Score (QMG, **B**), Myasthenia Gravis Composite Score (MGC, **C**) during 12 months of follow-up for MG-001 participants who received six once-weekly doses (n=7). **D:** Change from Baseline in MGC Score after initial dosing and retreatment in a participant experiencing relapse at Month 12. **E:** Relative change in serum anti-acetylcholine receptor antibody levels in the three participants with detectable antibodies at baseline. Each line represents one patient.

Three of the 4 participants with a history of anti-AChR antibody had detectable levels at baseline, and all three showed reductions in antibody levels by Month 6 (−17%, -44%, and -65%). These reductions continued at Month 9 (−35%, -100% [undetectable], and -70%), and persisted at Month 12 (−33%, -44% and -65% reductions, **Figure 1E**). The participant whose anti-AChR antibody was undetectable at Month 9 and -44% at Month 12 also had worsening disease from Month 9 to Month 12 but did not opt for retreatment. Five of the 7 participants had circulating anti-meningococcal antibodies at baseline, all from prior vaccination. As we previously reported, there was a small but detectable decrease in these antibodies at Month 3.^1^ The reduction deepened by Month 9 (−48% [-25% to -75%]) and stabilized by Month 12 to lower but still protective levels (−48% [-35% to -69%], **Figure s2**), following a similar pattern to the anti-AChR antibody reduction. In contrast, there was no appreciable decrease in total immunoglobulin levels (**Figure s3**), including total IgG levels at Month 3 (−21% [-56% to +14]).

In summary, we observed continued clinical improvement and autoantibody reductions after BCMA-directed rCAR-T treatment that persisted through the one-year follow-up period. A patient with clinical relapse at Month 12 again achieved MSE after redosing. The favorable safety profile of this mRNA CAR-T treatment is in contrast to DNA-based CAR-T treatments, which require lymphodepletion chemotherapy with potential hematologic toxicities and oncogenic risk from genomic integration of CAR DNA. DNA-based CAR-T therapy also targets a much broader population of non-pathogenic CD19-positive cells, which can cause unnecessary immunosuppression.^8^ Therapies targeting BCMA, but not CD19 or CD20, have advantageous effects on patients’ autoantibody signatures.^9^ Our observations suggest that using rCAR-T therapy to target BCMA-positive cells—including LLPCs—can result in durable depletion of autoantibodies and clinically meaningful improvement in MG severity scores without severe toxicity, agammaglobulinemia, or increased risk of infection. These data support continued development of Descartes-08 in myasthenia gravis and other autoantibody-associated autoimmune disorders.

## Data Availability

Access to anonymised, individual, and trial-level data (analysis datasets) will be provided by request from qualified researchers performing independent, rigorous research, after review and approval of a research proposal and statistical analysis plan and execution of a data sharing agreement.

## Contributors statement

All authors had full access to the study design information, reviewed, edited, and provided final approval of the manuscript content, and had final responsibility for the decision to submit for publication. Study design: MK, MDM, TM. Investigation and data collection: NC, GS, MHF, TV, JFH, TM. Data analysis: MDM, CAS. Data verification: MK, JFH. Data interpretation: CAS, CJ, MK, MDM, TV, TM, JFH. Writing — original draft: MDM.

## Acknowledgments

We thank all the study participants and trial teams, as well as members of the Study Monitoring Committee: Gil Wolfe, Syed Abbas Ali, and Mihriye Mete. Research reported in this publication was supported by the National Institute of Neurological Disorders and Stroke of the National Institutes of Health under Awards Number R25NS088248 and NS115426-01A1. The content is solely the responsibility of the authors and does not necessarily represent the official views of the National Institutes of Health. MG-001 study was sponsored by Cartesian Therapeutics, Inc.

---

**Figure s1.**
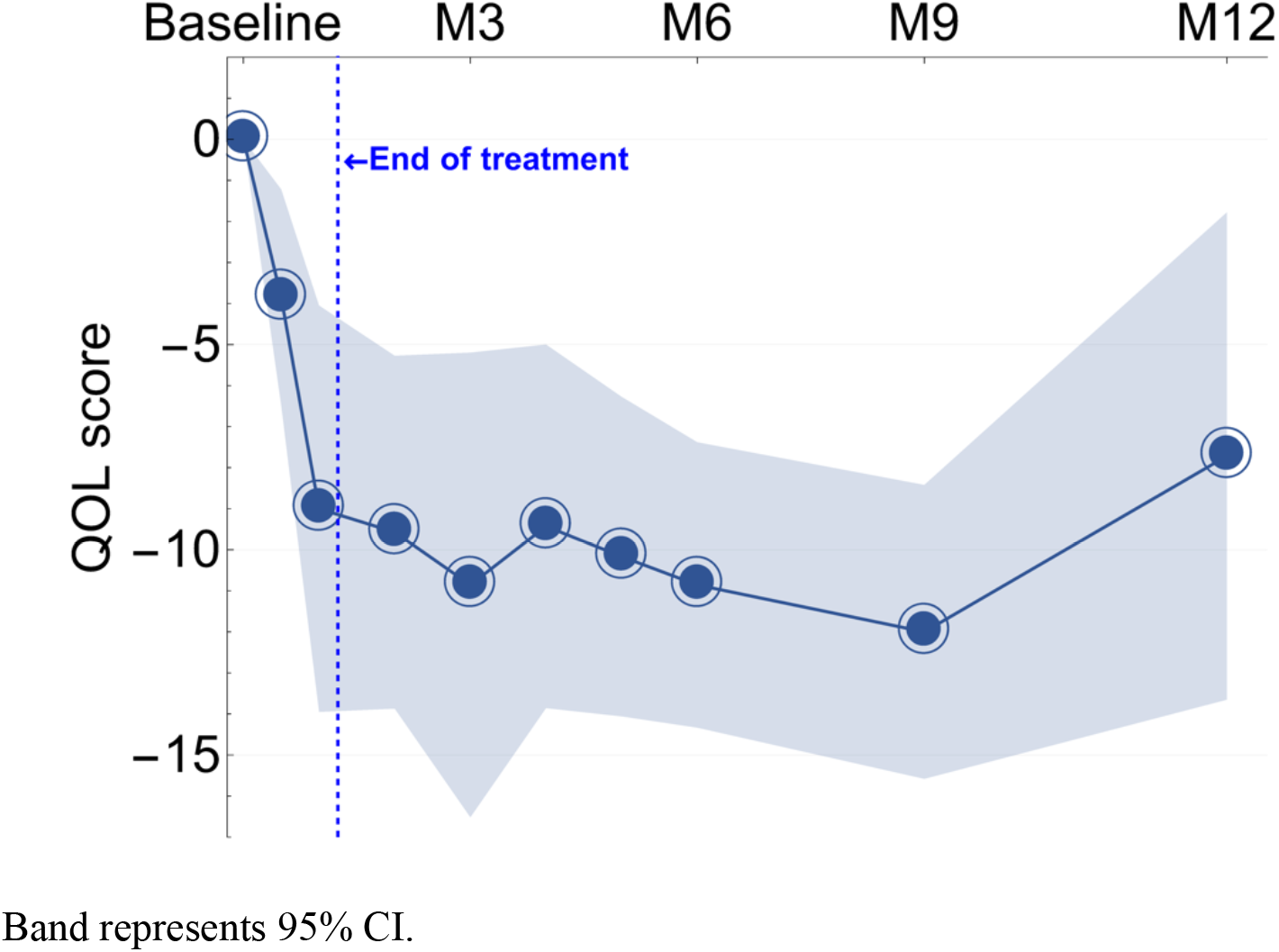
Mean change from baseline in Myasthenia Gravis Quality-of-Life Revised Scale (MG-QoL-15r) score. Band represents 95% CI.

**Figure s2.**
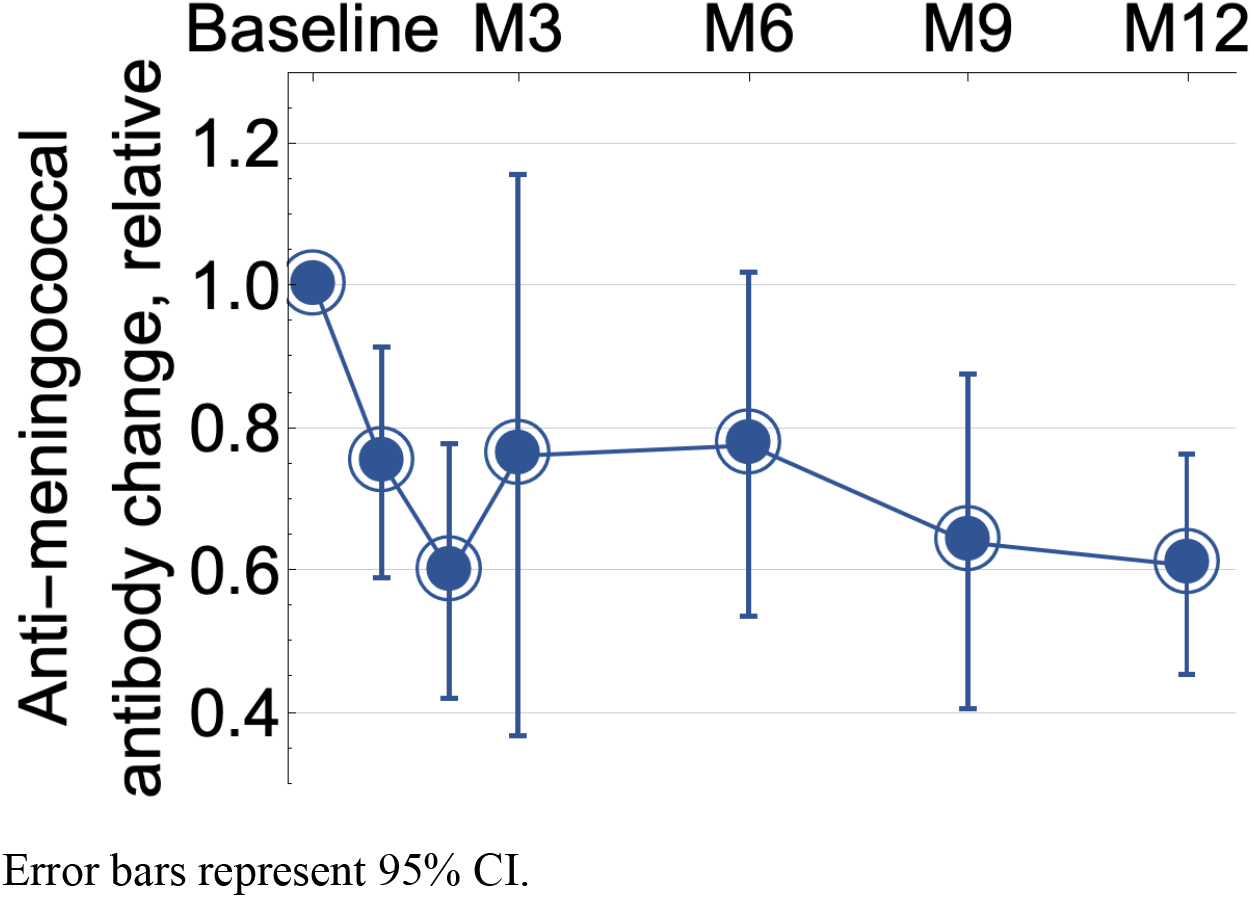
Relative change from baseline in anti-meningococcal antibody titers (all serogroups). Error bars represent 95% CI.

**Figure s3.**
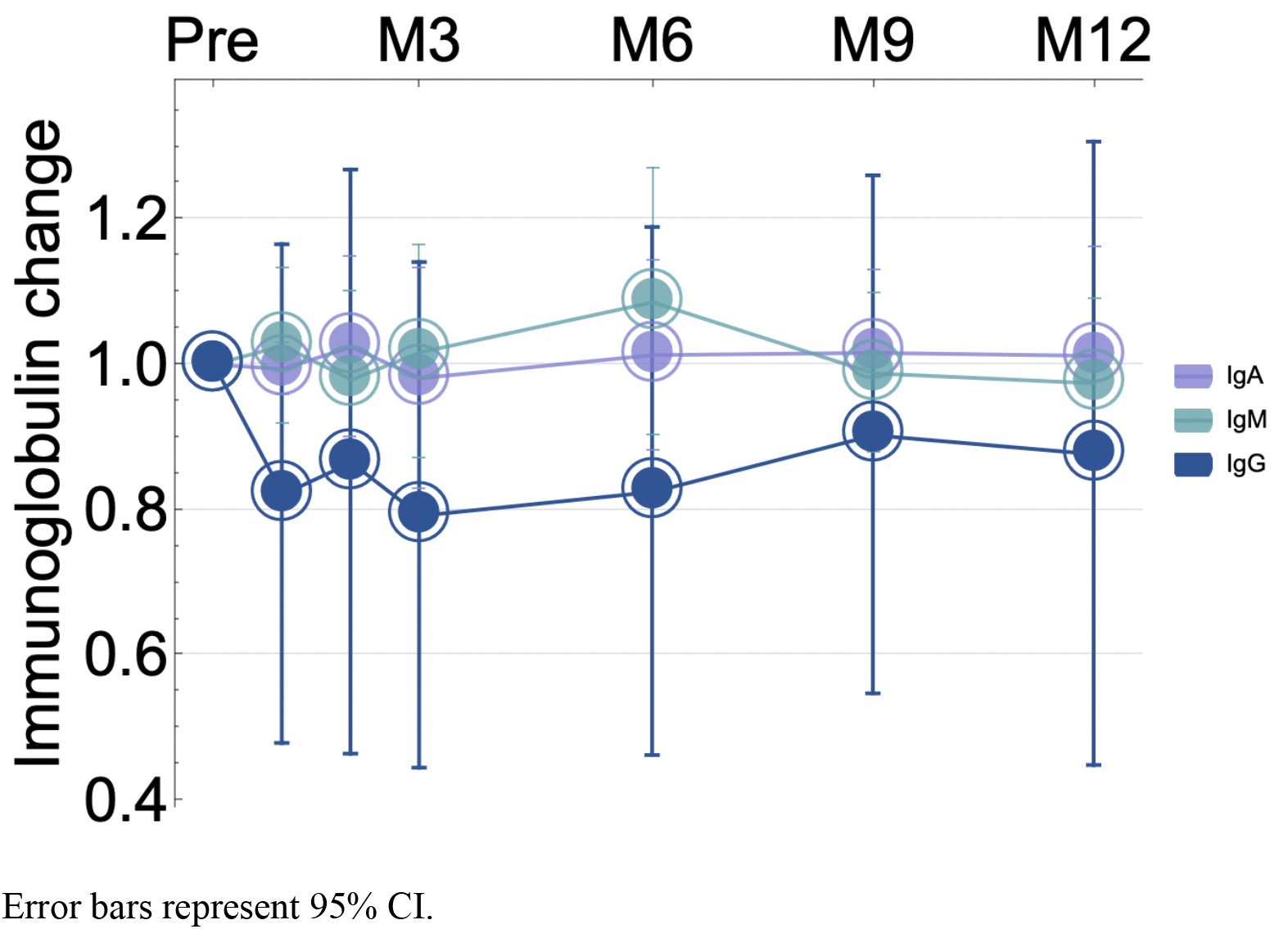
Relative change from baseline in total immunoglobulin levels. Error bars represent 95% CI.

## References

1 Granit V, Benatar M, Kurtoglu M, et al. Safety and clinical activity of autologous RNA chimeric antigen receptor T-cell therapy in myasthenia gravis (MG-001): a prospective, multicentre, open-label, non-randomised phase 1b/2a study. Lancet Neurol 2023; 22: 578–90.

2 Lin L, Cho S-F, Xing L, et al. Preclinical evaluation of CD8+ anti-BCMA mRNA CAR T cells for treatment of multiple myeloma. Leukemia 2021; 35: 752–63.

3 Vissing J, Jacob S, Fujita KP, et al. ‘Minimal symptom expression’ in patients with acetylcholine receptor antibody-positive refractory generalized myasthenia gravis treated with eculizumab. J Neurol 2020; 267: 1991–2001.

4 Muppidi S, Silvestri NJ, Tan R, Riggs K, Leighton T, Phillips GA. Utilization of MG-ADL in myasthenia gravis clinical research and care. Muscle Nerve 2022; 65: 630–9.

5 Burns TM, Conaway M, Sanders DB, MG Composite and MG-QOL15 Study Group. The MG Composite: A valid and reliable outcome measure for myasthenia gravis. Neurology 2010; 74: 1434–40.

6 Barnett C, Katzberg H, Nabavi M, Bril V. The quantitative myasthenia gravis score: comparison with clinical, electrophysiological, and laboratory markers. J Clin Neuromuscul Dis 2012; 13: 201–5.

7 Burns TM, Sadjadi R, Utsugisawa K, et al. International clinimetric evaluation of the MG-QOL15, resulting in slight revision and subsequent validation of the MG-QOL15r. Muscle Nerve 2016; 54: 1015–22.

8 Haghikia A, Hegelmaier T, Wolleschak D, et al. Anti-CD19 CAR T cells for refractory myasthenia gravis. Lancet Neurol 2023; 22: 1104–5.

9 Bodansky A, Yu DJ, Rallistan A, et al. Unveiling the autoreactome: Proteome-wide immunological fingerprints reveal the promise of plasma cell depleting therapy. MedRxiv 2023.12.19.23300188.

